# Correlation analysis of heart rate variations and glucose fluctuations during sleep

**DOI:** 10.1101/2023.07.20.23292857

**Authors:** Taira Kajisa, Toshiya Kuroi, Hiroyuki Hara, Toshiyuki Sakai

## Abstract

**Objective:** The body’s glucose concentration is influenced by carbohydrate intake, insulin-induced carbohydrate reduction, and hepatic glycogen breakdown induced by stress hormones. This study investigated the potential of employing glucose fluctuations as a measure of stress by examining the relationship between heart rate variability (HRV) data and glucose levels during sleep in healthy subjects.

**Methods:** In this cross-sectional study, a chest-worn electrocardiogram (ECG) and continuous glucose monitoring device (CGM) were respectively used to monitor the heart rate intervals and glucose fluctuations of five subjects (two males, three females) during sleep. A time-series correlation analysis was performed on the HRV data extracted from heart rate intervals and the corresponding glucose fluctuation data.

**Results:** The time-series analysis of ECG and CGM data collected from subjects during sleep (n = 25 nights) revealed a moderate negative correlation between glucose levels and HRV, with a cross-correlation coefficient of r = –0.453.

**Conclusion:** Similar to HRV, changes in stress levels can be detected by observing glucose fluctuations, particularly during sleep when the impact of food intake can be eliminated. Our findings highlight a significant correlation between glucose levels and HRV, indicating that glucose fluctuations can be used as an indicator of autonomic nervous system activity.

## INTRODUCTION

Sleep is pivotal for maintaining overall health as diminished sleep quality increases the risk of lifestyle diseases, including diabetes, myocardial infarction, and stroke [1–5]. Moreover, studies have suggested that diminished sleep quality could be used in the early detection of mental disorders [6,7]. According to recent reports, approximately one-third of American adults sleep for less than the recommended 7 h, with 50–70 million individuals afflicted by sleep disorders [8,9]. Furthermore, studies have found that daytime stress is associated with degraded sleep quality [10,11] and that the use of digital devices, including computers and smartphones, affects sleep quality [12,13]. Thus, the precise monitoring, evaluation, and analysis of daily sleep quality are paramount.

Several studies have used biosensing data gathered during sleep to assess sleep quality. Primarily, sleep studies have employed electrophysiological measurements, specifically electroencephalograms (EEG), gathered using scalp electrodes. Various EEG frequency bands are analyzed to monitor rapid eye movement (REM) sleep stages, non-REM sleep, and non-REM sleep sub-stages (N1, N2, and slow-wave sleep) [14,15]. In addition to identifying relaxation, awakening, and sleep cycles from EEG brainwave frequency patterns, several methods have been developed to evaluate sleep based on heart rate and heart rate variability (HRV) data from electrocardiograms (ECG). Previous studies have reported a correlation between ECG and EEG measurements in evaluating sleep quality [16,17]. With the advent of wearable devices (e.g., chest bands and wristwatch heart rate monitors), accurate measurements of heartbeat intervals have been validated against conventional ECGs [18]. For example, chest-worn ECGs have been used to analyze HRV in evaluating autonomic nervous system (ANS) activity [19]. Existing methodologies mainly involve external monitoring using wearable devices. Thus, there is a growing demand for more convenient, high-precision devices to monitor ANS activity.

Given this background, we hypothesized that direct measurement of biomarkers within the body would allow for more precise monitoring of ANS activity. We considered biomarkers related to sympathetic nervous system activity (e.g., epinephrine, cortisol, and glucagon) and those related to parasympathetic nervous system activity (e.g., serotonin and dopamine). However, these biomarkers are present in extremely dilute concentrations in body fluids, thus requiring large experimental equipment including a high-performance liquid chromatography system for measurement [20–22]. Although several kits used to quantify cortisol from saliva are available, biosensors that can directly sense other stress and relaxation hormone biomarkers are lacking because of their extremely dilute concentrations in body fluids. Consequently, EEG and HRV have been chosen as surrogate biomarkers. However, in this study, blood glucose fluctuations, influenced by stress hormones, were considered as a novel biomarker that can reflect sympathetic nervous system activity [23–26]. While glucose fluctuations primarily result from oral carbohydrate intake, the secretion of stress hormones because of a surge in sympathetic nervous system activity can elevate blood glucose levels (Figure 1). Thus, we hypothesized that blood glucose fluctuations during sleep could be used to reflect pure changes in sympathetic nervous system activity, uninfluenced by oral glucose intake. Continuous glucose monitoring (CGM) systems facilitate regular interstitial glucose monitoring at few-minute intervals. Previous studies using CGM data during sleep in diabetic patients suggested an association between glycemic control and sleep alterations [27]. Consequently, glucose analysis during sleep could be used to measure sympathetic nervous system activity and sleep quality. This study aims to examine the changes reflective of stress during sleep by comparing and conducting a time-series analysis of glucose fluctuations and HRV during sleep in healthy subjects.

**Figure 1.**
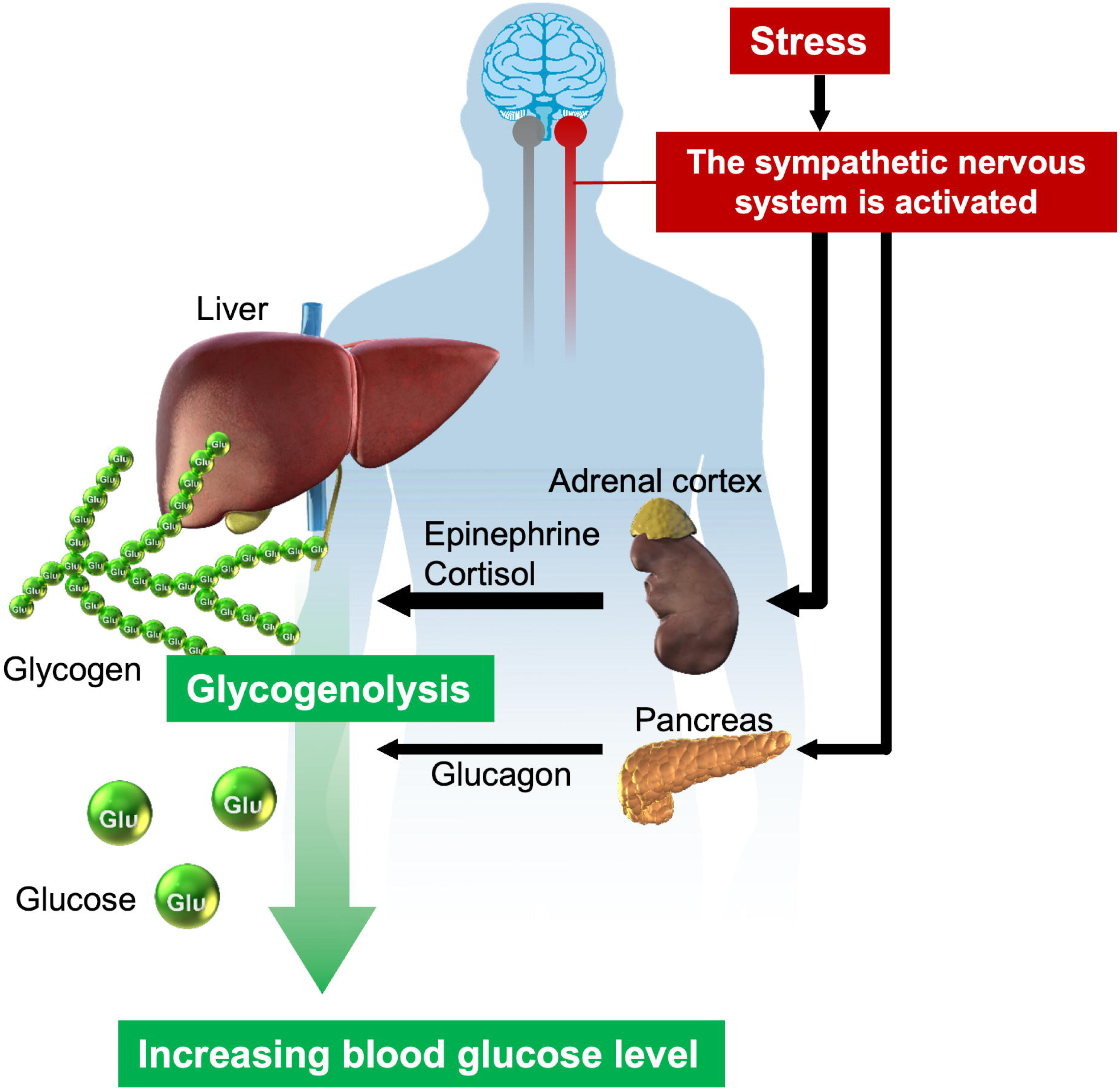
Schematic illustrating the mechanism of increased blood glucose levels during sympathetic nervous system dominance induced by stress.

## EXPERIMENTAL SECTION

### 2.1. Subjects and Materials

This study was approved by the Ethics Review for Medical Research Involving Human Subjects at Toyo University (Approval Number TU2021-006-TU2022-H-012). Prior to study initiation, participants were comprehensively briefed about the study’s objectives, methodology, and potential risks. Informed consent was obtained from all participants after providing a consent form that detailed the experimental protocol and data collection procedures. All participants were healthy adults, aged ≥20 years, who were selected based on health examinations indicating fasting blood sugar levels ≤100 mg/dL, HbA1c levels ≤6.0%, and metabolic syndrome assessments falling below the standard threshold. CGM devices (FreeStyle Libre, Abbott, USA) were used to perform temporal measurements of interstitial fluid glucose levels. Data were collected via the LibreLink app on smartphones. In adherence to data privacy regulations, data were procured from LibreView (https://www.libreview.com/) for analysis. A Polar H10 Heart Rate Sensor (Polar Electro Oy, Finland) chest strap electrocardiograph and Polar Pacer (Polar Electro Oy) wristwatch were used to conduct heart rate interval measurements. Both devices collected data at the same sampling frequency (1,000 Hz).

### 2.2. Study Protocol

The primary objective of this study was to elucidate the correlation between temporal changes in glucose levels and heart rate intervals. CGM and heart rate interval data were acquired during subjects’ bedtime and from ECG, respectively. Five healthy participants (two males and three females) wore a CGM sensor while performing normal activities to monitor temporal glucose data. After participants had their meals 3 h prior to bedtime, they wore an ECG sensor and wristwatch and began measurements upon retiring to sleep. The measurements concluded upon awakening and the removal of the ECG sensor. In addition to the correlation analysis between glucose levels and heart rate intervals, CGM data were collected before and after events expected to induce changes in ANS activity. CGM data were gathered during a five-day period from a long-distance runner (in his 20s): four days before a major race. Furthermore, CGM data were gathered before and after a sauna session from one participant (in his 40s).

### 2.3. Statistical Analysis

Fundamental HRV analysis was conducted using ECG-derived R-wave interval data. Time-domain measures were used to calculate the average heart rate (HR), average RR interval (RRi), coefficient of variation of RR intervals (CV_R-R_), root mean square of successive differences of RR intervals (RMSSD), and percentage of heartbeats with consecutive RR interval differences exceeding 50 ms (pNN50). The ratio of the low-frequency component (LF) to the high-frequency component (HF) of HR interval data was presented as a frequency-domain measure, reflecting the balance between sympathetic and parasympathetic nerves. HR intervals were smoothed using a moving average coefficient of *k* = 25 to demonstrate trend variability. A Fourier transform model of the power spectral density of R-wave interval data was used to calculate the LF/HF ratio. For the collected CGM data, basic values such as 24-h average glucose, sleep time average glucose, and daytime average glucose were calculated for each participant. The Pearson product-moment correlation coefficient was used to analyze the correlation between glucose variability observed through CGM and HRV observed through ECG. The lag between glucose variability and HRV was considered in the correlation analysis to obtain the cross-correlation coefficient. The temporal lag of the two time-series data was set from −20 to 20, and the correlation coefficient for each lag was calculated. All statistical calculations and time-series analysis were performed using Excel 2016 and Python 3.0.

## RESULTS

### 3.1. Data Acquisition from Participants During Sleep

HR interval and glucose data were collected from five healthy participants during sleep. Data were selected based on interviews and glucose peaks. The data collected during sleep on days without an oral glucose peak within 3 h before bedtime were chosen. Table 1 shows the parameters related to body composition, quantity of data from each participant during sleep (total n = 25), and average sleep measurement time. The average age of participants was 40.6±8.6 years, and participants had an average body composition of 20.7±3.9, which denoted a standard body type according to WHO body mass index (BMI) percentile definitions. Sleep time (defined as the duration in which the chest band was worn) and HR measurements were conducted and averaged at 7 h and 18 min±1 h and 23 min.

**Table 1.** Characteristics of study participants.

| Variables | Participants (n = 25) |
| --- | --- |
| Age at test, Mean±SD | 40.6 ± 8.6 |
| Number (male/female) | 5 (2/3) |
| BMI (kg/m<sup>2</sup>), Mean±SD | 20.7 ± 3.9 |
| Average bedtime (hours : minutes), Mean±SD | 7:18 ± 1:23 |

### 3.2. Heart Rate Interval Measurement Using ECG

HR intervals were measured from participants during sleep using a chest band-type electrocardiograph and wristwatch-type measuring device. The data were sourced from five days of sleep data for each participant. For each participant, the average HR, average RRi, RMSSD, CV_R-R_, and pNN50 were calculated as time-domain indicators during sleep for HRV analysis. The LF/HF ratio was calculated as a frequency-domain indicator. Figure 2 displays the data of each participant to illustrate the variations, while Table 2 presents the collective statistical data (n = 25). The heart rate and heart rate interval during sleep were 69.8±14.2 bpm and 890±241, respectively.

**Figure 2.**
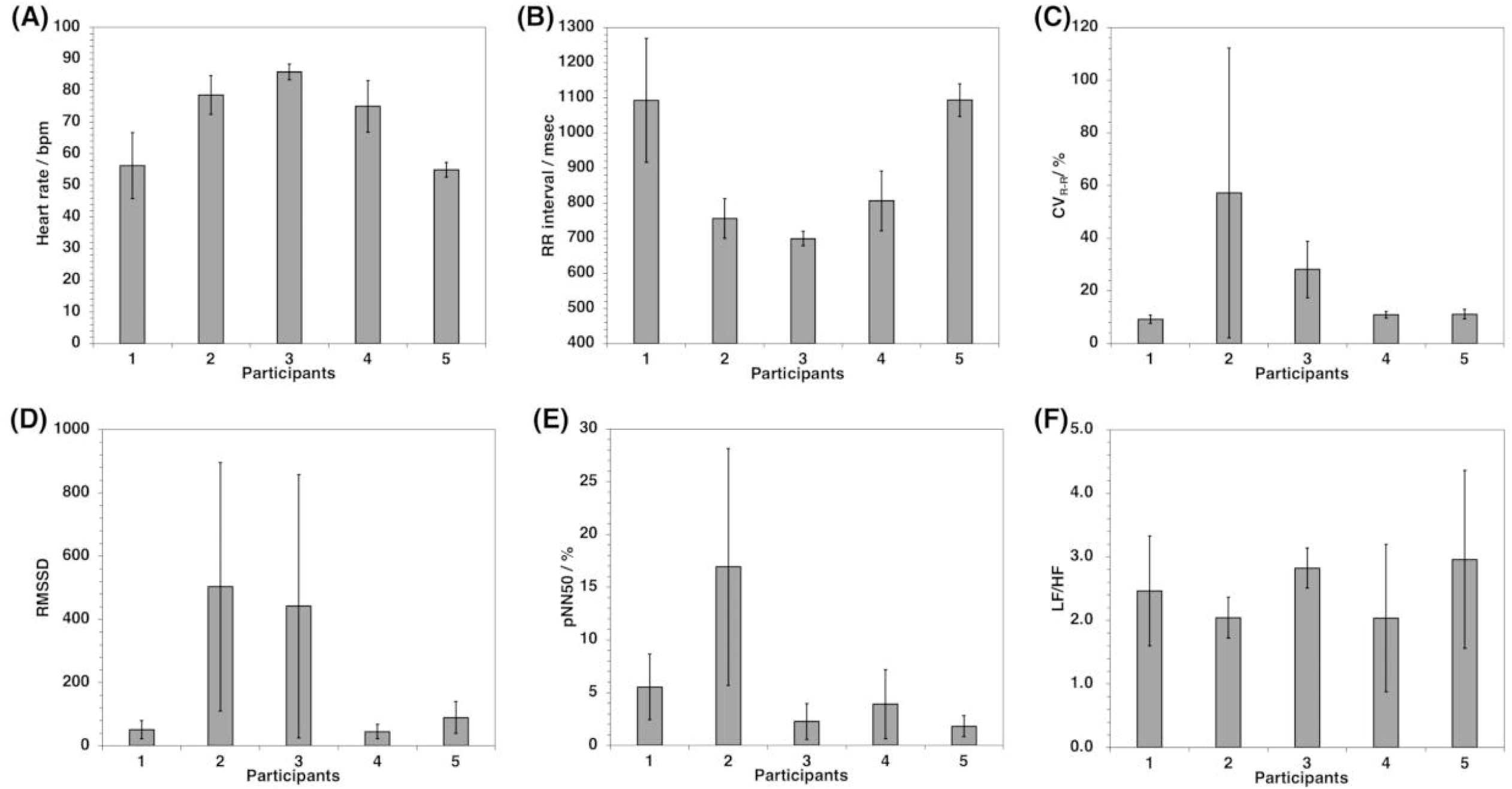
Comparative diagram of parameters related to heart rate variability (HRV) derived from sleep electrocardiogram data among subjects (n = 5): A) heart rate (HR), B) RR interval (RRi), C) coefficient of variation of RR intervals (CV_R-R_), D) root mean square of successive differences of RR intervals (RMSSD), E) percentage of heartbeats with consecutive RR interval differences exceeding 50 ms (pNN50), and F) ratio of low-frequency to high-frequency power in HRV (LF/HF). RMSSD and pNN50 are indices of parasympathetic nervous system activity.

**Table 2.** Time-domain parameters of heart rate variability in a sample population (n = 25)

| Time domain parameters | Values (n = 25) |
| --- | --- |
| HRV(bpm), mean ± SD | 69.8 ± 14.2 |
| RRi (msec), mean ± SD | 890 ± 241 |
| RMSSD, mean ± SD | 226 ± 226 |
| CV<sub>R-R</sub> (%), mean ± SD | 23.3 ± 20.4 |
| pNN50 (%), mean ± SD | 6.09 ± 6.23 |
| LF/HF, mean ± SD | 2.46 ± 0.93 |

### 3.3. Temporal Glucose Measurement Using CGM

Table 3 shows the average glucose values for each participant during the test and sleep. The average CGM glucose value for one day was 102.0±7.6, whereas the average glucose value during sleep was 90.9±6.0. The glucose values ranged from 70 mg/dL to 180 mg/dL for 78.9% and 86.6% of the time throughout the day and during sleep, respectively. Figure 3 shows the glucose data of participants for one day and during sleep. For all participants, the average glucose value during sleep was 5.8–15.4 mg/dL (mean: 11.1 mg/dL) lower than that for the day. As can be inferred from the data in Table 3 and Figure 3, no significant difference was observed in glucose values among participants.

**Figure 3.**
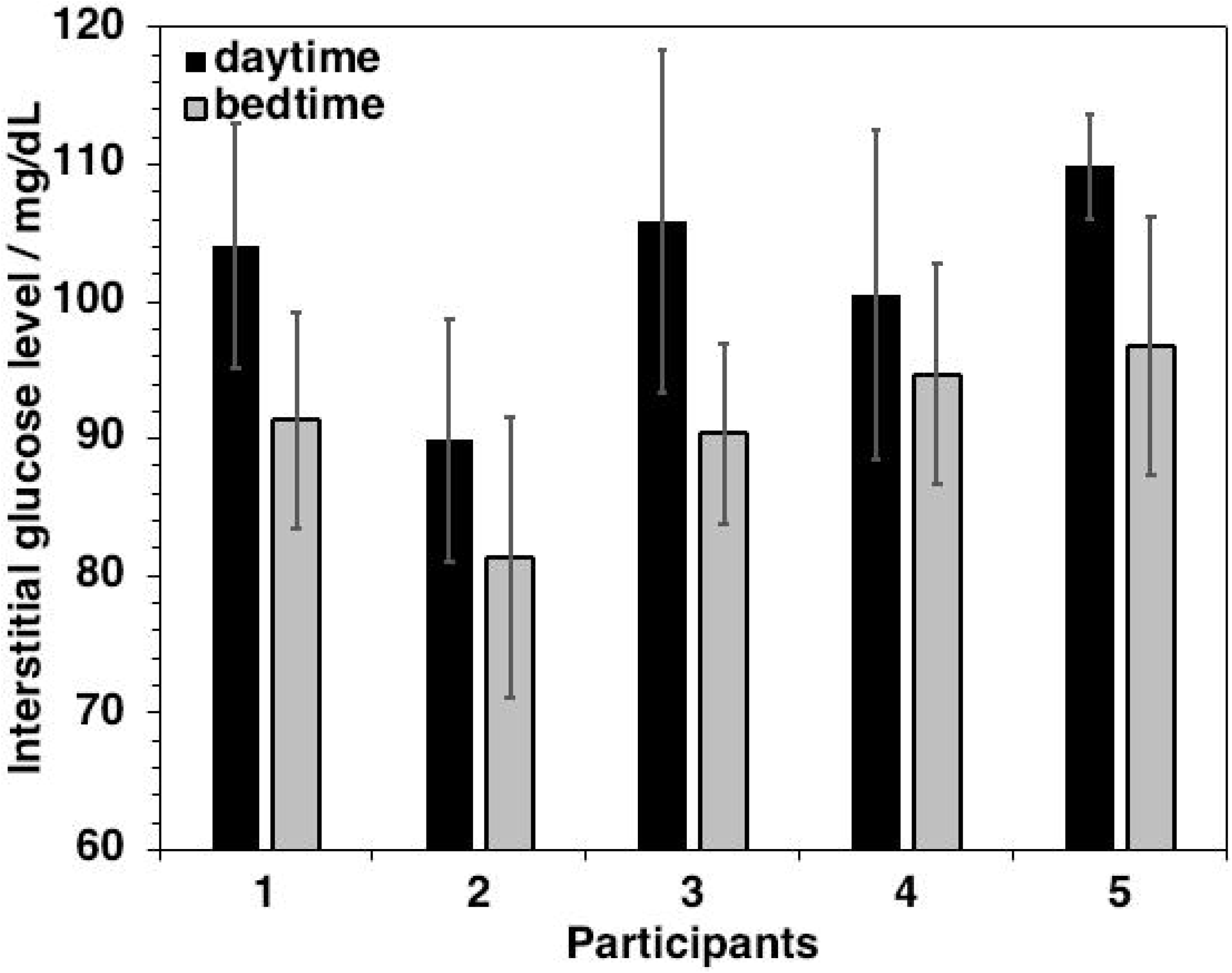
Interstitial glucose levels during sleep and wakefulness among subjects.

**Table 3.** Comparison of interstitial glucose levels between 24 hours and bedtime (n=25) based on the range from international consensus.

| Interstitial glucose data (n = 25) | 24 hours | Bedtime |
| --- | --- | --- |
| Average glucose level (mg/dL), mean ± SD | 102.0 ± 7.6 | 90.9 ± 6.0 |
| Time >180 mg/dL (%), mean ± SD | 4.6 ± 8.0 | 0.0 ± 0.0 |
| Time 70–180 mg/dL (%), mean ± SD | 78.9 ± 22.0 | 86.6 ± 18.7 |
| Time <70 mg/dL (%), mean ± SD | 16.5 ± 22.8 | 13.4 ± 19.3 |

### 3.4. Time-Series Data Analysis of Heart Rate Interval and Glucose Value

Temporal changes in heart rate interval and glucose fluctuations from five participants during sleep were provided for time-series analysis (n = 25). The results are shown in Figures 4 and 5. Figure 4 shows the temporal changes in graphs and cross-correlations of each time-series analysis from relatively small data of the cross-correlation function (r = −0.741) to large data (−0.328) in the n = 25 data. As indicated in Figures 4A–C, an increase in the heart rate interval was associated with a decrease in glucose fluctuation, and vice versa. Additionally, Figures 4D– F illustrate the offset of the cross-correlation of heart rate interval and glucose fluctuation data, as per Pearson’s product-moment correlation function. When the lag was set to ±20 (ca. ±40 min) and the lag of the cross-correlation coefficient was analyzed, a negative cross-correlation existed within the range of lag ±20. Figure 5 presents the analysis of the variations of the cross-correlation function between each participant. The average value of the cross-correlation function of time-series data of glucose and heart rate interval fluctuations in all data was −0.453±0.089. The lag of both data averaged at −1.08 (SD: 12.4).

**Figure 4.**
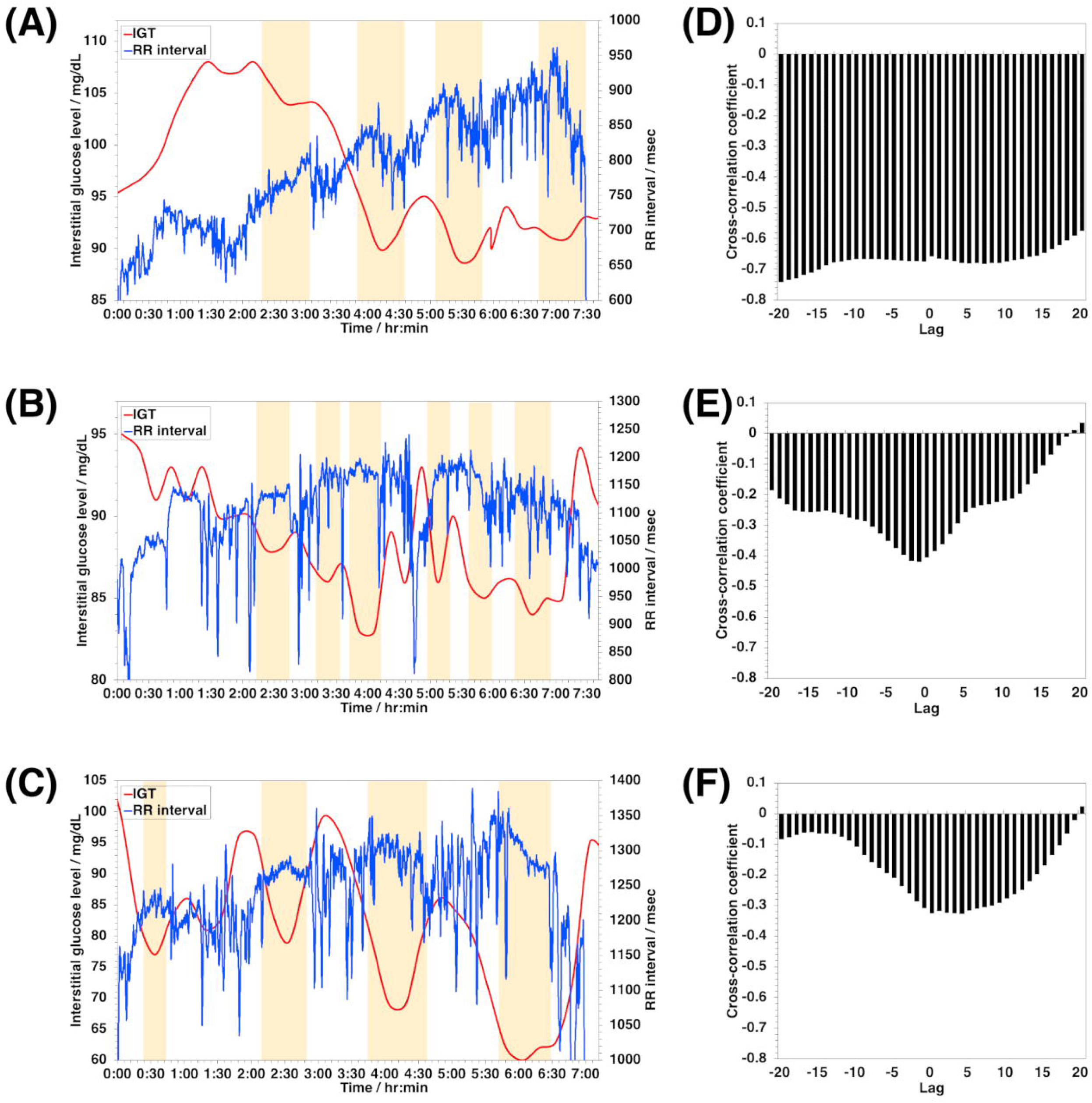
Temporal data of heart rate variability and glucose fluctuations during sleep: A–C) Interstitial glucose level (IGL, red) and RR interval (blue). D–F) Correlation coefficient data considering the time lag between heart rate intervals and glucose fluctuations.

**Figure 5.**
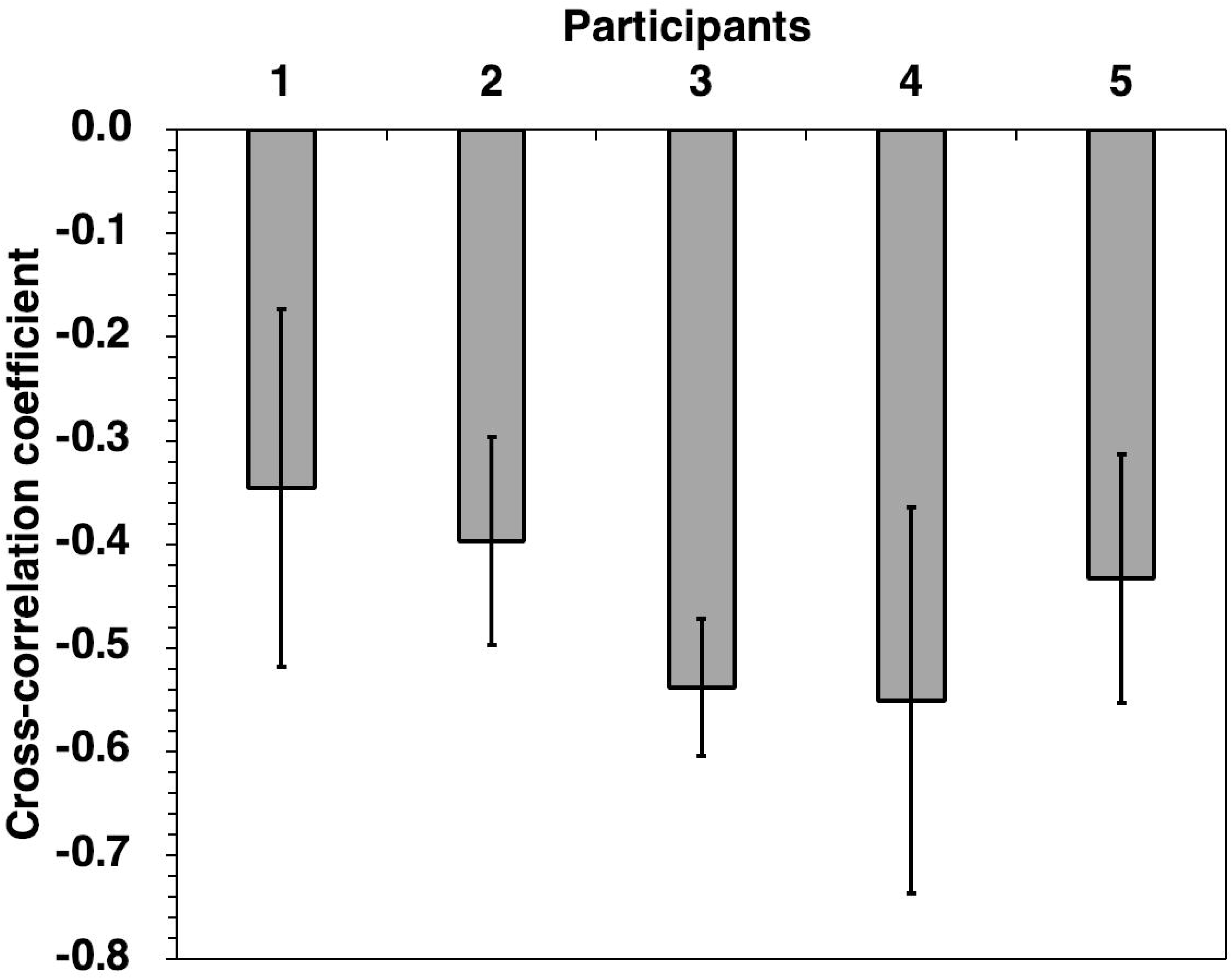
Correlation coefficients of heart rate variability and glucose fluctuations during sleep among subjects.

### 3.5. Glucose Data During Sleep Before and After the Event

Glucose fluctuations during sleep before and after significant events were examined. In Figure 6A, glucose data during sleep four days before a major race of a long-distance runner were compared. The average glucose value during sleep from 0:00 to 5:00 was 84.8 ± 5.5 mg/dL at 2– 4 days before the race and 104.7 ± 3.7 mg/dL the day before the race (p < 0.001). Additionally, Figure 6B presents the glucose data of a male participant (in his 40s) during sleep before and after a sauna session. When comparing glucose values from 2:00 to 8:30 (average sleep time), the average glucose value during sleep was 103.3 ± 10.6 mg/dL at five days before the sauna and 83.6 ± 6.6 mg/dL after the sauna (p < 0.001). Significant differences in glucose fluctuations during sleep were observed in both events.

**Figure 6.**
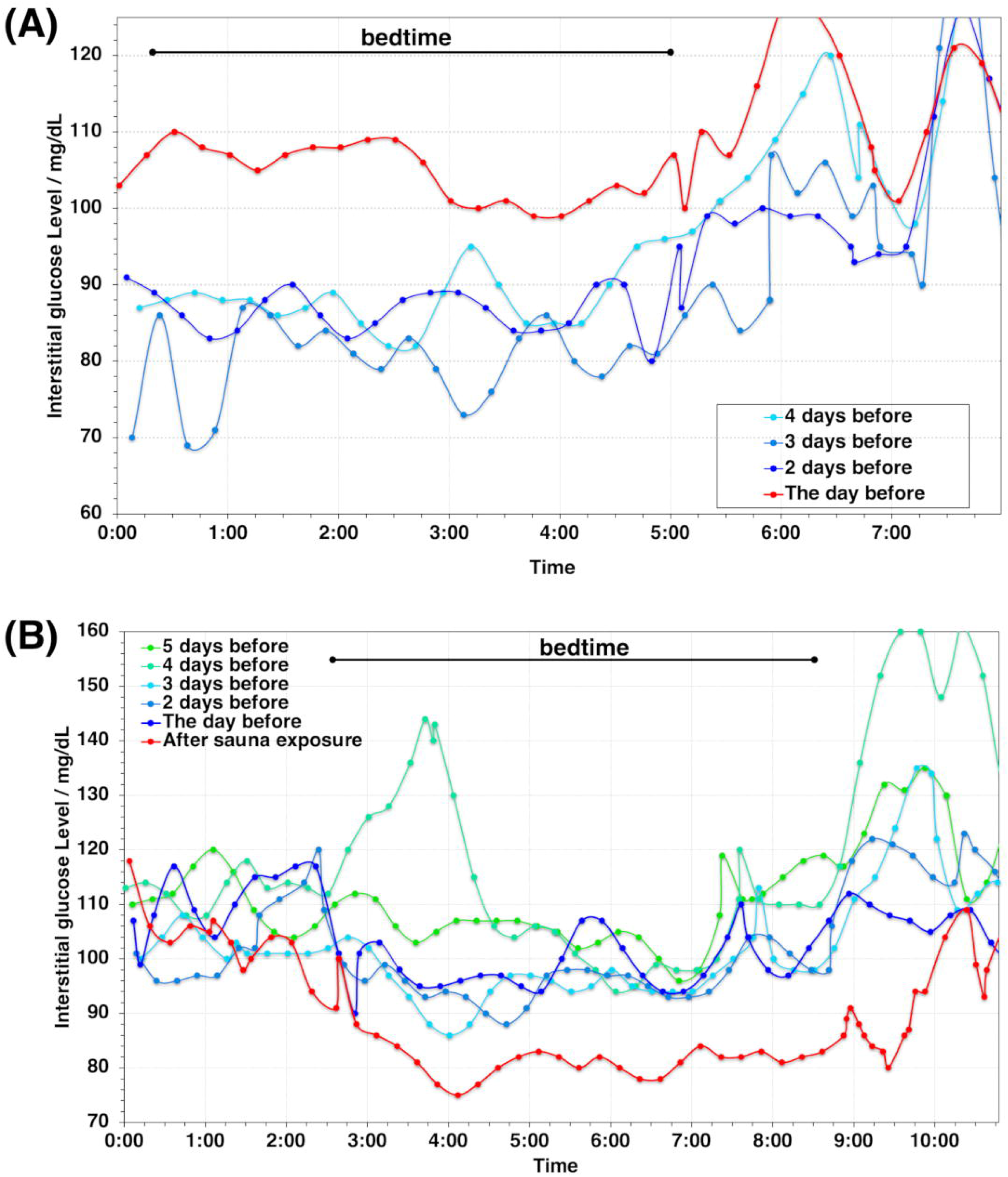
Temporal data of interstitial glucose levels during sleep: A) from four days before and the day after a major race in a long-distance runner; B) before and after sauna exposure in a subject in his 40s.

## DISCUSSION

The present study explored the possibility of using glucose fluctuations measured by CGM to monitor stress. The FreeStyle Libre CGM sensor, which monitors temporal glucose from interstitial fluid based on the principle of electrical current measurement with enzymatic methodology, has shown a significant correlation with blood glucose levels [28–31]. Our approach consisted of comparing glucose fluctuations with ECG data. Previous studies have confirmed the effectiveness of ECG in demonstrating a correlation with traditional stress analysis methods, such as EEG. While the accurate measurement of HRV with ECG necessitates a resting monitoring environment, glucose monitoring with CGM exhibits fluctuations because of oral glucose intake and insulin secretion. The secretion of stress hormones, a response to sympathetic nervous system activity, triggers the breakdown of liver glycogen, causing a subsequent rise in blood glucose levels. Considering these dynamics, we selected sleep data for comparison to minimize the influence of noise from both devices and conduct a robust correlation analysis. Previous reports have focused on deriving sleep cycles from HRV during sleep for stress monitoring. Simultaneously, some studies have reported glucose fluctuations during sleep using CGM, primarily for managing blood glucose during sleep in patients with type I and II diabetes [27,32]. In our study, the subjects were individuals with non-metabolic syndrome (BMI: 20.7 ± 3.9, Table 1) and normal daily glucose values (Table 3). According to the guidance on blood glucose control evaluation targets for adults, the indicators for target glucose range, time below target glucose range, and time above target glucose range confirm a low risk for type I and II diabetes [33]. Hence, we performed a correlation analysis of HRV and glucose fluctuations during sleep in healthy adults with an average age of 40.6 years (SD: 8.6). HRV analysis was conducted using heart rate intervals obtained from ECG. As suggested by the RMSSD, an indicator of parasympathetic nervous system activity in the time-domain indicators for heart rate variability, sympathetic nervous system activity was not significantly stressed during the entire sleep time (Figure 2 and Table 2). Moreover, the LF/HF ratio (a frequency-domain indicator) was consistent with values reported for healthy individuals, suggesting little deviation from past HRV analysis results [34–36]. Additionally, chronoamperometric glucose monitoring using CGM revealed that no subjects had values indicative of type I or II diabetes (as depicted in Figure 3 and average values in Table 3), confirming the healthy status of our participants. Notably, the average glucose level during sleep was on average 11.1 mg/dL lower than the average glucose level for the day. This difference is likely because of the influence of meals and daily life stress on blood glucose levels, implying that such impacts are minimized during sleep. Based on these findings, we subjected the HRV analysis by ECG and glucose analysis by CGM to time-series analysis using Pearson’s product-moment correlation coefficient. Given that ECG-based HRV follows temporal values in near real time, while glucose measurements by CGM experience a time lag in response to sympathetic nervous system activity and a time delay between blood and interstitial glucose levels, we accounted for this lag by using a cross-correlation function (Equation 1):

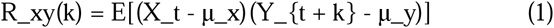

where R_xy(k) represents the cross-correlation of X and Y at lag k; E[.] denotes the expectation (average); X_t is the value of X at time point t; Y_{t + k} is the value of Y at time point t + k; and μ_x and μ_y are the average values of X and Y, respectively. A correlation analysis of heart rate variations and glucose fluctuations during sleep was conducted in the present study. As shown in Figure 4, the results indicated a negative correlation between glucose levels and HRV, suggesting that an increase in glucose was associated with a decrease in HRV, and vice versa.

The observed correlation occurred within a time lag of approximately ±40 min, implying the possibility of a time delay between glucose fluctuations in the interstitial fluid and sympathetic nerve activity reflecting hepatic glucose metabolism, as well as the infiltration of glucose from the vasculature into the interstitial fluid. Previous studies have suggested time lags of 8–40 min between glucose secretion and metabolism, as well as the influx of glucose into the interstitial fluid. Based on the findings of this analysis, the correlation coefficient between HRV and interstitial glucose fluctuations in the study participants ranged from −0.345 to −0.551, with a mean of −0.453±0.089, indicating a moderate negative correlation (Fig. 5). Studies have previously examined the role of blood glucose control on cardiac autonomic function measured by HRV in patients with type 1 diabetes, suggesting a correlation between blood glucose fluctuations and cardiac autonomic function. In this study, a correlation between HRV and glucose fluctuations during sleep was demonstrated in healthy individuals, suggesting that glucose fluctuation data can be used for analyzing stress with higher accuracy and reflecting autonomic function. Additionally, Figure 6 presents two notable examples illustrating the association between glucose fluctuations during sleep and autonomic function. Figure 6A displays the glucose fluctuations during sleep of a long-distance runner from three days before to the day prior to a major race. As can be seen, glucose levels during sleep on the day prior to the race were consistently higher by an average of 10 mg/dL compared with the two preceding days. This observation suggested the potential dominance of sympathetic activity associated with autonomic regulation during sleep on the day prior to the competition, assuming that noise from meals and exercise could be ignored. Conversely, Figure 6B compares glucose fluctuations during sleep on the day of sauna exposure with those on other days, revealing significantly lower glucose levels during sleep immediately following sauna exposure. Previous reports have indicated that acute sauna exposure leads to parasympathetic dominance and a decrease in sympathetic activity of the cardiac autonomic system [37]. The observed lower glucose levels during sleep after sauna exposure may suggest the suppression of hepatic glucose breakdown mediated by stress hormones because of the parasympathetic predominance induced by sauna exposure.

In summary, this study revealed a negative correlation between HRV measured using a simple chest band and glucose fluctuations measured using CGM during sleep. The findings indicate that changes in glucose levels can be used to analyze stress and evaluate sleep quality in healthy individuals, which can be used for pre-diagnostic medical applications to prevent future diseases.

## CONCLUSION

The present study investigated the relationship between glucose fluctuations measured by CGM and sympathetic nervous system activity during sleep. The findings demonstrated that HRV is correlated with glucose fluctuations, suggesting a potential link between ANS function and glucose metabolism. This correlation highlights the importance of considering physiological and biochemical factors in understanding sleep-related changes in glucose regulation. Furthermore, the analysis revealed that sleep duration and quality play a crucial role in maintaining optimal metabolic function. Sleep disturbances, such as insomnia and sleep apnea, are associated with increased glucose fluctuations and impaired HRV, indicating a potential pathway through which sleep disorders may contribute to metabolic dysregulation. The results emphasize the importance of promoting healthy sleep habits and addressing sleep disorders as part of comprehensive metabolic health management. Moreover, the study demonstrated the feasibility and efficacy of CGM in monitoring glucose fluctuations during sleep. CGM provides valuable insights into the dynamics of glucose levels, revealing the impact of meals, stress, and sympathetic nervous system activity on overnight glucose regulation. This information can be used to create personalized approaches to glucose management, particularly in individuals with diabetes. Overall, the findings highlight the intricate interplay between sleep, glucose metabolism, and sympathetic nervous system activity. Future research should focus on elucidating the underlying mechanisms and developing targeted interventions to optimize sleep quality, promote metabolic health, and reduce the risk of associated health conditions. Overall well-being and health outcomes can be improved by understanding and addressing these factors.

## Data Availability

All data produced in the present study are available upon reasonable request to the authors.

https://doi.org/10.6084/m9.figshare.23707884

## AUTHOR INFORMATION

### Notes

The authors declare no competing financial interest.

## ACKNOWLEDGEMENT

Part of this work was supported by HAKUJU INSTITUTE FOR HEALTH SCIENCE Co., Ltd. The authors wish to thank all the healthy subjects who participated in the project.

